# Centroid to SUVpeak distance is a prognostic PET biomarker in high-grade glioma

**DOI:** 10.1101/2025.08.22.25334230

**Authors:** Jesús J. Bosque, David Molina-García, Julián Pérez-Beteta, Ana M. García Vicente, Víctor M. Pérez-García

**Affiliations:** Departamento de Matemática Aplicada, Escuela Técnica Superior de Ingeniería Agronómica, Alimentaria y de Biosistemas (ETSIAAB), Universidad Politécnica de Madrid (UPM), Madrid, Spain; Mathematical Oncology Laboratory (MOLAB), University of Castilla-La Mancha (UCLM), Spain; Department of Mathematics, Facultad de Educación, University of Castilla-La Mancha (UCLM), Ciudad Real, Spain; Instituto de Investigación Sanitaria de Castilla-La Mancha (IDISCAM), Spain; Department of Mathematics, Escuela Técnica Superior de Ingenieros Agrónomos, University of Castilla-La Mancha (UCLM), Ciudad Real, Spain; Nuclear Medicine Department, Complejo Hospitalario Universitario de Toledo, Toledo, Spain; Department of Mathematics, Escuela Técnica Superior de Ingeniería Industrial, University of Castilla-La Mancha (UCLM), Ciudad Real, Spain

**Keywords:** SUVpeak, normalized distance from hotspot to centroid (NHOC), glioma, PET, survival, prognostic biomarke, ^18^F-fluorocholine

## Abstract

**OBJECTIVE:** The normalised distance from the metabolic hotspot to the tumour centroid (NHOC) in Positron Emission Tomography (PET) scans is an imaging biomarker previously shown to have prognostic value in non-small cell lung cancer (NSCLC) and breast cancer (BC). The primary objective of this study was to assess whether this metric could also possess prognostic value in high-grade glioma patients.

**METHODS:** We retrospectively collected ^18^F-FCHOL diagnostic PET studies from 61 patients with confirmed high-grade glioma. We delineated the metabolically active tumour regions and calculated the geometrical tumour centroid. SUVpeak was obtained and its distance to the geometrical centroid calculated and normalised by the mean spherical radius—a linear measure of tumour size. We used this metric (NHOCpeak) and SUVpeak to perform Kaplan–Meier survival analysis and multivariate analyses.

**RESULTS:** NHOCpeak (log-rank p-value = 0.02) and SUVpeak (p-value < 0.001) were uncorrelated (Spearman’s *ρ* = 0.094) and separated patients in groups with different median survivals (10.1 and 7.7 months respectively). Taking NHOCpeak and SUVpeak as independent variables for the location and activity in the hotspot, we separated the patient cohort into 4 groups (high/low NHOCpeak and high/low SUVpeak). Patients with low SUVpeak and low NHOCpeak—the most beneficial group—had the longest survival, with a median benefit of 16.4 months (log-rank p-value = 0.017) compared to the second-best (high NHOCpeak/low SUVpeak). Patients with low NHOCpeak and low SUVpeak had a median survival gain of 21.9 months (log-rank p-value < 0.001) with respect to the group with the worst outcome (high SUVpeak/high NHOCpeak).

**CONCLUSIONS:** The metric NHOCpeak, calculated as the relative distance between the hotspot of activity and the tumour centroid normalised by tumour size, predicts survival in high-grade glioma imaged with ^18^F-FCHOL complementing other PET-based prognostic biomarkers.

## INTRODUCTION

Gliomas are primary brain tumours with high morbidity and mortality for which few novel therapeutic alternatives have emerged in the last decades. In particular, high-grade gliomas (WHO grades III and IV), comprising anaplastic astrocytoma and oligodendroglioma, as well as glioblastoma, are very aggressive and most often recur after surgery plus adjuvant therapy (*1*). Patient segmentation and treatment personalisation is important to improve survival in these malignancies. Given these challenges, imaging biomarkers may help stratify patients for treatment and follow-up.

Positron emission tomography (PET) imaging has been used as a means to extract prognostic biomarkers in many malignancies (*2–4*). Values extracted from tumour images such as the metabolic tumour volume (MTV), total lesion activity (TLA) or the maximum of the so-called standardised uptake value (SUVmax) have been related to survival in different cohorts. Another simple classic biomarker is SUVpeak, which measures the maximum activity within a volume of interest by averaging the activity of neighbouring voxels to provide a more robust metric. In neuro-oncology in particular there are suitable radiotracers in use, such as 6-^18^F-fluoro-L-DOPA (^18^F-FDOPA) (*5,6*) or ^18^F-fluorocholine (^18^F-FCHOL) (*7*). PET studies with these tracers have led to the definition of imaging biomarkers in glioma (*8–10*).

PET images provide a macroscopic visualization of the underlying metabolic activity exerted by tumour cells and provide access to spatially structured information that can be further exploited for patient prognosis. PET imaging biomarkers inspired by mathematical models have gained momentum in the last decade. For instance, so-called scaling laws relate a theoretical TLA to MTV, thus providing a reference to which the actual in activity can be compared to define the distance to the scaling law (DSL) which bears prognostic value in different malignancies (*11,12*).

In that context, the normalised distance between the hotspot of activity—either measured by the SUVmax or SUVpeak—and the tumour centroid (NHOC) has been proposed as a biomarker for relapse and survival (*13*). The metric, normalised by the mean spherical radius (MSR), that is the radius of a sphere with the same volume as the tumour, gives a value that locates the hotspot of activity between the centre of the tumour (NHOC=0) or towards its edge (NHOC near 1). Mathematical models suggested that more evolved tumours would show activity shifted towards their edge and, therefore, would show a higher NHOC.

NHOCmax, which uses the distance between the centroid and the voxel with SUVmax, was initially proven to have prognostic value in cohorts of breast cancer and resectable non-small cell lung cancer (NSCLC) imaged with ^18^F-fluorodeoxyglucose (^18^F-FDG) (*13*). Later, its use was extended to ^18^F-FDG imaged NSCLC with the voxel with SUVpeak, instead of the SUVmax, as a proxy for the location of the metabolic hotspot, giving rise to NHOCpeak (*14*). Hovhannisyan-Baghdasarian et al. validated the use of NHOC in two cohorts of advanced NSCLC patients (*15*). Hong et al. demonstrated the value of NHOC to predict response to neoadjuvant chemotherapy in a group of 135 patients with breast cancer imaged with ^18^F-FDG (*16*). The biomarker has recently also been added to the calculation software LIFEx (*17*). Finally, Zheng et al. used the normalised distance from the SUVmax to perimeter (NHOPmax) to discriminate survival in a cohort of 164 patients with operable stage IA-IIIA lung adenocarcinoma imaged with ^18^F-FDG (*18*).

The use of NHOC as a prognostic biomarker has so far only been tested on breast cancer and NSCLC, however the fundamental nature of the modeling framework leading to its definition suggests that it could be applicable beyond those malignancies. Here we used a cohort of 61 patients with high-grade gliomas, imaged at diagnosis using ^18^F-FCHOL PET, to investigate whether the location of the hotspot identified by the NHOC provides relevant information about patient prognosis.

## MATERIALS AND METHODS

### Patients

Patients were enrolled in a prospective non-randomised multicentre study (FuMeGA: Functional and Metabolic Glioma Analysis) with approval from a local Ethical Committee (internal code B/176/2016). All patients signed written informed consent for the use of their anonymized medical data. Patients suspected of having glioma were subjected to a basal ^18^F-FCHOL positron emission tomography/computed tomography (PET/CT) scans after magnetic resonance imaging (MRI). A cohort of 61 patients with histologically confirmed high-grade glioma and an operable lesion were included. After surgery, patients underwent adjuvant standard treatment depending on clinical status: radiotherapy combined with temozolomide plus temozolomide and/or second-line chemotherapy agents versus palliative support.

### Image acquisition

PET/CT scans were performed on a Discovery DSTE-16s (GE Medical Systems) scanner in three-dimensional (3D) mode. The acquisition started 40 minutes after intravenous administration of 185 MBq of ^18^F-FCHOL. A low-dose CT transmission study (modulated 120 kV and 80 mA) was first performed without injected contrast, to later continue with the 3D emission study over 20 minutes of acquisition time. The voxel size in the resulting PET images was 2.3 × 2.3 × 3.3 millimetres, in a 128 × 128 matrix reconstructed using the CT images in an iterative reconstruction algorithm for attenuation correction.

MRI scans were contrast enhanced T1-weighted sequences with gradient-echo sequences and volumetric spoiled gradient recalled echo or volumetric fast field echo without magnetisation transfer after intravenous administration of a single dose of gadobenate dimeglumine (0.1 mmol/kg) with a delay of 6-8 minutes. Images were then acquired in the axial plane using a 1.5 T or 3 T MR imaging unit. Images were delineated and processed, and their representative values (total tumour volume, necrotic volume, width of contrast enhancing rim, tumour surface and tumour regularity) were calculated as described in previous works (*19–21*).

### SUVmax calculation

From the raw image data, counts were transformed to SUV in each voxel by the formula

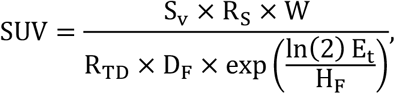

with *S*_*v*_ being the counts stored value, R_S_ the rescale slope, W the patient weight, R_TD_ the radiopharmaceutical injected dose, *D*_*F*_ its decay factor, and *H*_*F*_ its half-life. Finally, the elapsed time between injection and acquisition was considered by the factor E_t_. The resulting output was a spatially structured 3D matrix (image) containing the SUV values at each voxel.

### Tumour delineation

PET images in DICOM format were imported into the scientific software MATLAB and delineated semi-automatically using an image segmentation procedure with an in-house designed software. Only PET scans with a SUVmax larger than twice the background activity were considered. A nuclear medicine physician (A.M.G.V.) and an image engineer (J.P.-B.), both with more than 10 years of experience in tumour segmentation, manually selected the tumour. Subsequently, a grey-level threshold was used to select the metabolic tumour volume. Each slice of the image was then corrected manually to select the final extent of tumour activity, for instance, removing physiological activity contiguous to the tumour, such as the choroid plexus. See **FIGURE 1**(A) for visualisation.

**FIGURE 1.**
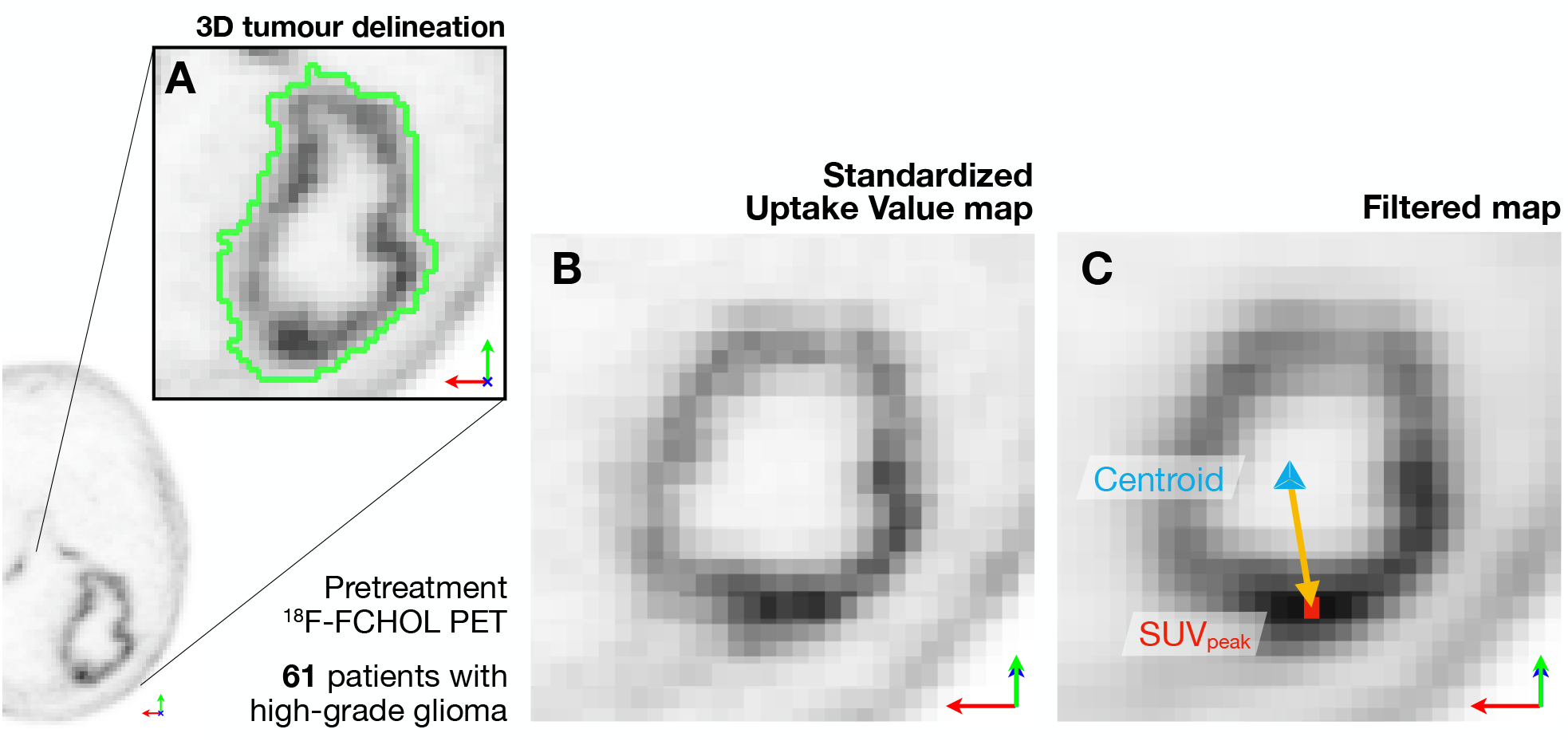
Research procedure. **(A)** PET scans were collected and the tumour delineated. In 3D SUV maps **(B)** we averaged every voxel’s neighbourhood activity. SUVpeak is the maximum activity in the filtered maps **(C)**. The distance between the tumour centroid and the voxel with SUVpeak normalised by a linear measure of volume (NHOCpeak) was used as a PET image survival biomarker. Notice the coordinate system is rotated in (B) and (C) as these show the plane in 3D that contains both the centroid and the SUVpeak. Positional arrows: green— anteroposterior axis, red—mediolateral axis, blue—craniocaudal axis.

### SUVpeak map calculation

The SUV map (**FIGURE 1**(B)) was spatially averaged at each voxel by applying a linear filter using a neighbourhood of 27 voxels surrounding each voxel (**FIGURE 1**(C)). The kernel of the linear filter was simply a 3×3 matrix containing the value 1/27 in every element. In practice, this means that for every voxel SUV activity was averaged in cubic regions of interest (ROI) of 0.47 cm^3^. This ROI was the most natural choice as it just represents the immediate neighbourhood for each voxel. The maximum metabolic activity in the filtered image is known as SUVpeak and indicates the region of maximum uptake in the filtered image, being thus less affected by noise than the SUVmax (*22*).

### Tumour volume

The volume of the metabolically active part of the delineated tumour was calculated as the number of voxels N multiplied by the volume of one voxel V_V_

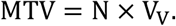

Voxels with lower metabolic activity enclosed within the tumour delineation for each axial slice were considered necrotic and their number N^*^ was considered for the calculation of the total tumour volume TTV

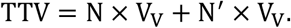

From the TTV, a linear measure of tumour volume was devised by considering the radius of a sphere with the same total tumour volume as

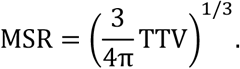

### Computation of NHOCmax and NHOCpeak

We calculated the position of the geometrical centroid of the tumour using its geometric definition, this is, averaging the values for all the N voxels of the metabolically active region of the segmented tumour in each of the spatial directions:

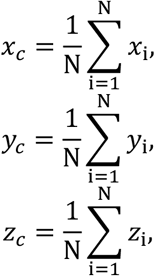

where *x, y, z* are the spatial coordinates at the centre of each voxel.

We then calculated the distance between the ^18^F-FCHOL hotspots (voxels with SUVmax and with SUVpeak, respectively) to the calculated tumour centroid by calculating the usual Euclidean distance between both points (see **FIGURE 1**(C) for graphical interpretation):

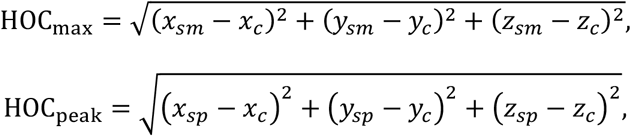

with (*x*_*sm*_, *y*_*sm*_, *z*_*sm*_) and (*x*_*sp*_, *y*_*sp*_, *z*_*sp*_) being the coordinates at the centre of the voxel with SUVmax and SUVpeak, respectively. Finally, we normalised these distances to obtain a size-independent measure by dividing by the *MSR*

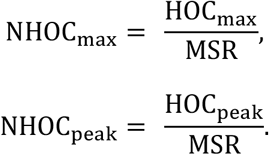

### Survival analysis

For each of the relevant metrics extracted from the image (SUVmax, SUVpeak, NHOCpeak, etc.) we performed survival analyses to discern whether the variables contained prognostic information and could therefore be used as prognosis image biomarkers. All analyses were made for overall survival (OS), i.e., the time from the date of the pre-treatment imaging study to the time of death or, in the case of censored patients, last follow-up examination.

For each of the analysed metrics we performed Kaplan–Meier survival analyses for OS, separating the cohort in two subgroups relative to the value that they have in the studied parameter. For each splitting threshold of the parameter, patients with a value lower than or equal to the threshold were included in one of the subgroups and their difference in survival was compared to the patients in the other subgroup with parameter higher than the threshold. We used a two-tailed test for the difference between survivals with a significance level of 0.05 for the p-values. Confidence intervals at a 95 % level were computed by Cox proportional hazards regression analysis. Every possible splitting threshold was tested, and the p-value obtained from the log-rank test used to define the significant difference in survival between subgroups.

Additionally, we computed Harrell’s concordance index score (c-index), which compares the survival of the two subgroups by considering all the possible combinations of individuals in the two groups and reporting the proportion of correct predictions made by the biomarker (*23*).

### Other statistical techniques

We used a Mann-Whitney U test to test difference in the number of local maxima using “ranksum” routine in MATLAB R2023b. To test whether dichotomic groups with different survival separated by a certain biomarker corresponded to different IDH groups we applied Fisher’s exact test to the contingency table with MATLAB’s “fishertest” routine. Differences between NHOCpeak (approximately normal) in IDH-WT and IDH-mut groups were analysed with Student’s t and Kolmogorov–Smirnov tests using “ttest2” and “kstest2”. Correlation between variables was assessed by Spearman’s correlation coefficient using “corr”. To perform multivariable Cox regression analysis we used IBM SPSS Statistics 30.0.0.0 (172).

## RESULTS

This study included a total of 61 participants, comprising 39 males and 22 females. The median age of the participants was 60.3 years (range: 23–80). Patient characteristics are summarised in **TABLE 1** according to the 2016 WHO classification. The diagnostic PET scans of each patient were analysed, and several measures characterizing the tumour’s metabolic state were calculated. For these values, including the statistical distribution of the markers, see **TABLE 2**.

**TABLE 1.** Patient characteristics.

| Characteristic | n (%) |
| --- | --- |
| Number of patients |  |
| Total | 61 |
| Male | 39 (64 %) |
| Female | 22 (36 %) |
| Age |  |
| Mean ± SD | 60.3 ± 13.6 |
| Range | 23–80 |
| High-grade glioma subtype |  |
| IDH-WT glioblastoma | 46 (75.4 %) |
| IDH-mut grade III astrocytoma | 8 (13.1 %) |
| IDH-mut grade IV astrocytoma | 3 (4.9 %) |
| Unknown IDH, grade IV | 3 (4.9 %) |
| Grade III oligodendroglioma | 1 (1.7 %) |
| WHO category |  |
| Grade III | 9 (15 %) |
| Grade IV | 52 (85 %) |

**TABLE 2.** Statistical distribution of PET features (61 patients)

| Variable | Median | Mean | SD | Range |
| --- | --- | --- | --- | --- |
| SUVmax | 3.52 | 3.91 | 2.02 | 0.55 – 9.73 |
| SUVpeak | 2.60 | 2.96 | 1.51 | 0.44 – 7.76 |
| NHOCmax | 0.71 | 0.71 | 0.22 | 0.36 – 1.40 |
| NHOCpeak | 0.62 | 0.65 | 0.25 | 0.13 – 1.41 |
| MTV (cm3) | 35.08 | 39.47 | 26.25 | 6.05 – 124.93 |
| TTV (cm3) | 36.02 | 42.81 | 30.24 | 6.05 – 142.50 |
| Total lesion activity | 40.31 | 52.74 | 43.06 | 1.57 – 196.86 |

### NHOCmax is a weak survival biomarker in gliomas

We started by studying the prognostic value of NHOCmax in high-grade glioma patients. For each patient, we measured their NHOCmax (see **FIGURE S1** for distribution) and performed Kaplan–Meier analysis for all possible thresholds. Although one level of NHOCmax threshold significantly separated the groups (NHOCmax = 0.585, log-rank p-value = 0.049, **FIGURE S2**), most thresholds were not significant, unlike previously published results on NSCLC and breast cancer (*13*).

To understand the difference between our glioma cohort and the NSCLC and breast cancer cases, we studied the distribution of local maxima in metabolic activity, that is, voxels with higher activity than their surroundings. We found that, while NSCLC and breast cancers tended to exhibit a single local maximum of activity and, occasionally, a small number of isolated hotspots, gliomas consistently showed a high number of hotspots (**FIGURE S3**). This reflects much higher heterogeneity in PET images of gliomas.

### NHOCpeak holds prognostic value for high-grade glioma patients

Given NHOCmax’s limited performance and the high spatial heterogeneity observed in gliomas, we explored whether SUVpeak-based NHOC (NHOCpeak) might better capture prognostic information. We considered the distance between the centroid and the voxel for SUVpeak and calculated the NHOCpeak for each patient (see **FIGURE S1** for histogram). Performing Kaplan–Meier analyses we found that NHOCpeak was able to significantly separate the cohort in groups of patients with different survival. For all NHOCpeak threshold values below 0.518, the group separation was found to be statistically significant (**FIGURE 2**(A)).

**FIGURE 2.**
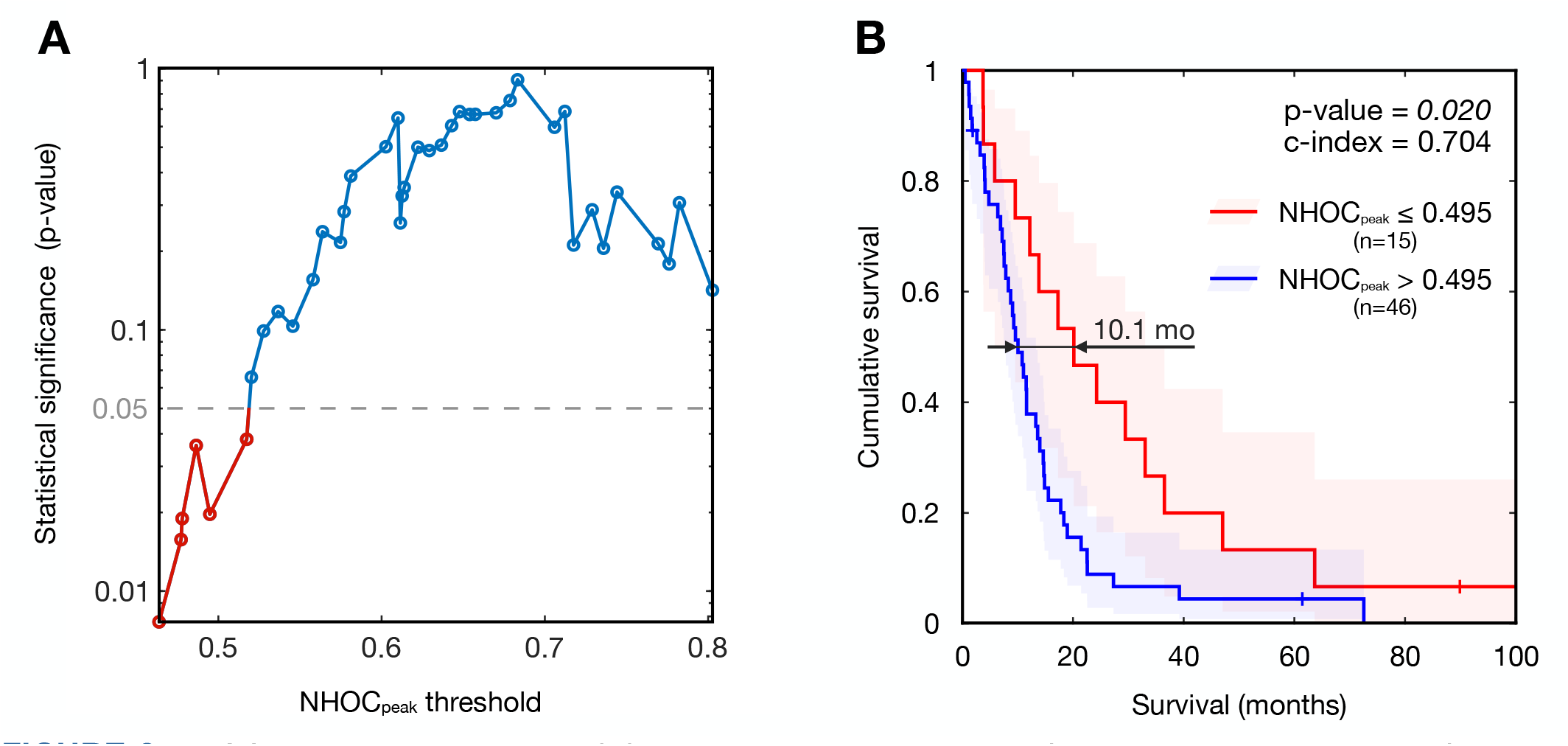
NHOCpeak survival analysis. **(A)** Log rank p-values obtained from the survival analyses of the two group separations by variable thresholds of the NHOCpeak. **(B)** Kaplan–Meier curve corresponding to the NHOCpeak that separates the patients’ groups with the best trade-off between significance level and balance between the number of patients in each group.

**FIGURE 3.**
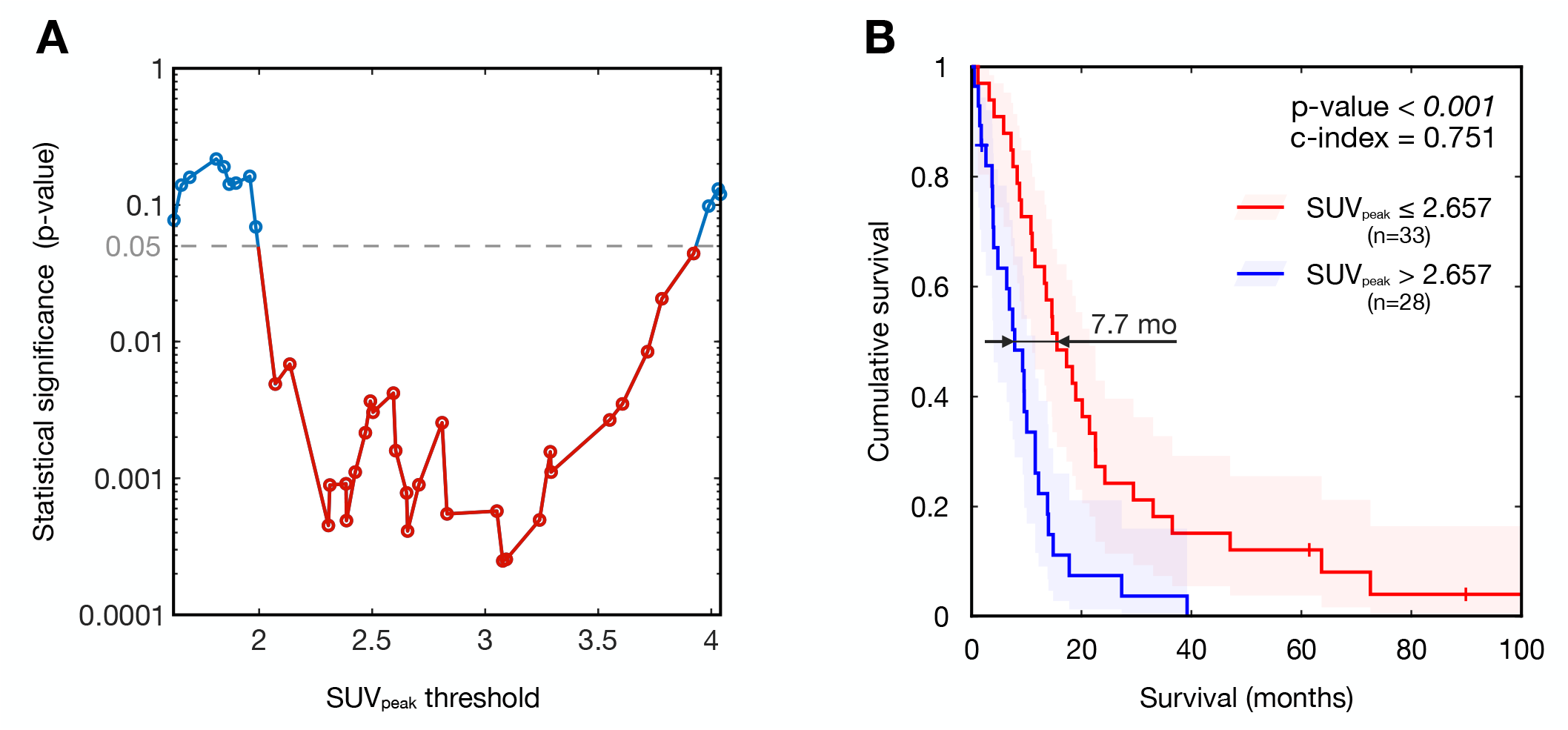
SUVpeak survival analysis. (A) Log rank p-values obtained from the survival analysis of the two group separations by variable thresholds (x axis). (B) Kaplan–Meier curve corresponding to the SUVpeak that separates the patient’s groups with the best equilibrium between significance level and equilibrium between patients in each group.

The best equilibrium between significance level and balance between groups appeared at the threshold NHOCpeak = 0.495. One quarter of the patients have a NHOCpeak less than or equal to that value. This subgroup has a significant increase in survival, with a median gain of 10.1 months (log-rank p-value = 0.02, c-index = 0.704, **FIGURE 2**(B)). Therefore, a lower value of NHOCpeak, corresponding to patients having peak metabolic activity close to the tumour centroid, was associated with increased survival in our glioma cohort.

### NHOCpeak does not differentiate IDH-WT from IDH-mut gliomas

Given that the cohort included both IDH-WT and IDH-mut patients, we investigated whether the long-survival group with low NHOCpeak could be predominantly composed of less aggressive IDH-mut patients. We created a contingency table (**TABLE S1**) according to IDH status (WT vs mut) and the corresponding NHOCpeak group (NHOC ≤ 0.495 vs NHOC > 0.495) considering the 58 patients with known IDH status. The proportion of IDH-mut patients in the low NHOCpeak group was 33 %, compared to 16 % in the high NHOCpeak group. However, Fisher’s exact test for the differences yielded non-significant result (p-value = 0.15), indicating that patients with low NHOCpeak are not significantly more likely to be IDH-mut than patients with high NHOCpeak. Coincidentally, IDH-WT and IDH-mut groups did not show significance differences in their NHOCpeak values (t-test p-value = 0.54, Kolmogorov– Smirnov test p-value = 0.25) as shown in **FIGURE S4**.

We then assessed whether NHOCpeak retained its predictive value in the group of 48 patients composed of uniquely IDH-WT high-grade gliomas. Survival analyses revealed that no NHOCpeak threshold was able to differentiate two groups with significantly different survival in this case (**FIGURE** S5). The lowest p-value in the log-rank test was 0.15. Therefore, for the subgroup formed by only IDH-WT gliomas, NHOCpeak was not able to discern survival.

### SUVpeak is also a survival biomarker

Since NHOCpeak and SUVpeak quantify complementary aspects of tumour metabolism (location vs. intensity), we next examined whether SUVpeak alone was also prognostic. We analysed the differences in survival between groups split by SUVpeak at successive thresholds and found that the metric significantly separated Kaplan–Meier curves across all thresholds between 1.98 and 3.92 (**FIGURE** 3(A)). Specifically, SUVpeak = 2.657 separates two well-balanced groups (n=33 for SUVpeak <= 2.657, and n=28 for SUVpeak > 2.657), with marked difference in survival (log rank p-value < 0.001). The median survival gain for the group with low SUVpeak group was 7.7 months. Therefore, both the location of the SUVpeak and its activity value are imaging biomarkers for high-grade glioma imaged with ^18^F-FCHOL PET. Moreover, the Spearman’s rank correlation coefficient between the two variables yielded a value of 0.094, indicating a lack of correlation.

We also tested whether these PET biomarkers were associated with previously reported MRI-derived biomarkers, namely, the total tumour volume, the volume of necrosis, the width of the contrast enhancing rim, tumour surface and tumour regularity as seen in MRI. We used Spearman’s rank correlation coefficient and found the values shown in **TABLE S2**, which reflect no correlation.

### Combination of NHOCpeak and SUVpeak distinguish survival differences

Both NHOCpeak and SUVpeak were independently associated with survival, although uncorrelated. We therefore assessed their combined prognostic value. **FIGURE 4**(A) shows a survival map, where the time to death of patients is indicated by the colour variable (for the 3 censored patients the colour indicates last follow-up). The map was split into four quadrants according to the relative value of SUVpeak and NHOCpeak: Group 1 (low SUVpeak, low NHOCpeak), Group 2 (low SUVpeak, high NHOCpeak), Group 3 (high SUVpeak, high NHOCpeak), and Group 4 (high SUVpeak, low NHOCpeak).

**FIGURE 4.**
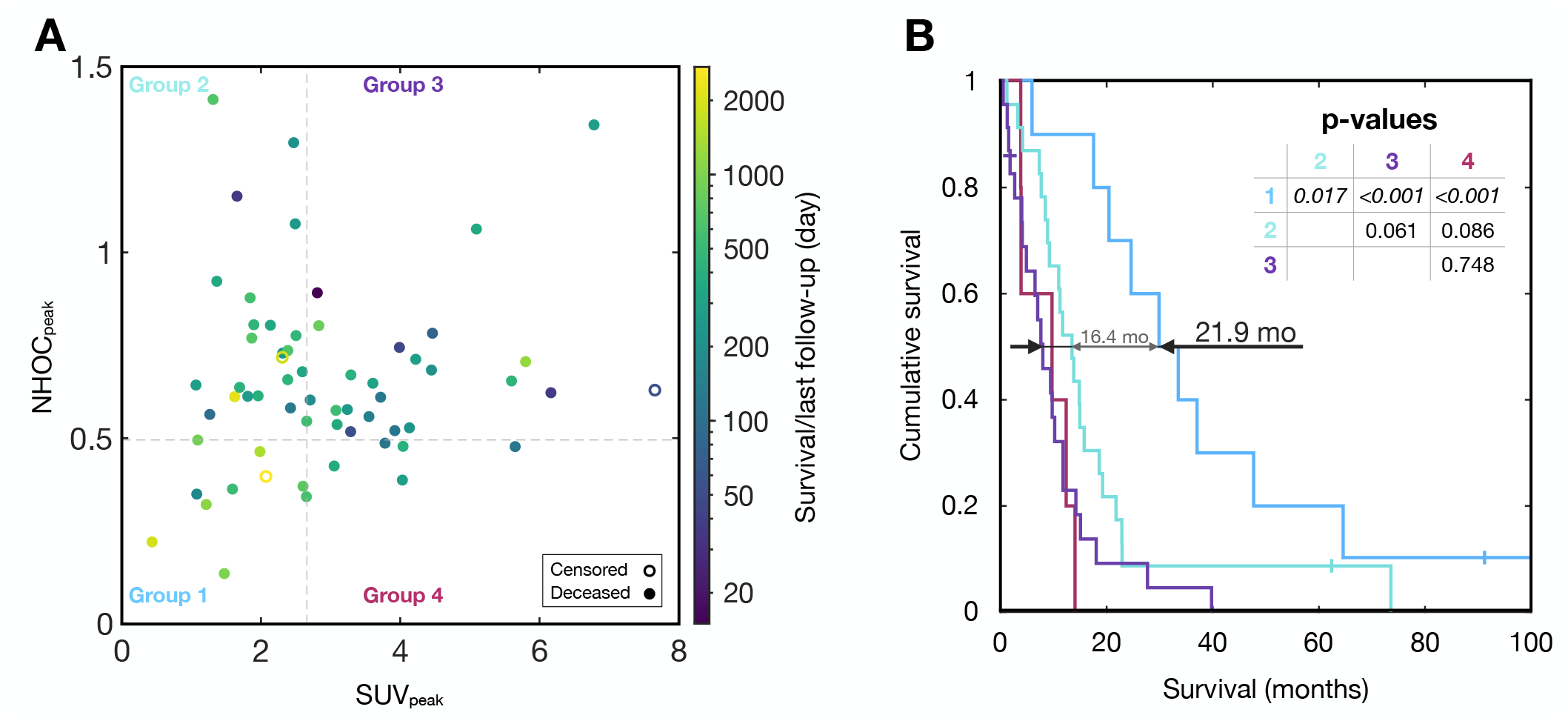
Results of the combination of SUVpeak and NHOCpeak. **(A)** Scatter plot of the SUVpeak (x axis) and NHOCpeak (y axis) associated to each patient. Each dot corresponds to one of the patients in the cohort. Closed points indicate deceased patients and open points, censored cases. The colour map indicates the survival time (closed points) or last follow-up time (open points). **(B)** Kaplan–Meier curves showing the survival of the 4 groups segmenting the patients in terms of their SUVpeak and NHOCpeak.

We also computed the Kaplan–Meier curves for these 4 groups (**FIGURE 4**(B)). Patients with low SUVpeak and low NHOCpeak (Group 1) showed a significantly better survival than all the other three groups. Group 1 showed a statistically significant difference (log-rank p-value 0.017) with the group with second best survival (Group 2, low SUVpeak/high NHOCpeak) and a median survival benefit of 16.4 months. Additionally, the median survival benefit of Group 1 with respect to the group with worst survival (Group 3, high SUVpeak/ high NHOCpeak) was as high as 21.9 months (p-value < 0.001).

Statistical significance for the difference in survival between Groups 2, 3 and 4 was weighed down by the low number of patients but it seems that patients in Group 2 have a better survival than those in Group 3 (log-rank p-value = 0.061). There were only five patients (8.2 %) in Group 4, which has a high SUVpeak while keeping NHOCpeak low. This suggests that, although high NHOCpeak does not directly imply high SUVpeak, patients with high SUVpeak have also a high NHOCpeak.

### Age is a prognostic biomarker independent of NHOCpeak

We found that age was also a prognosis biomarker in our cohort of high-grade glioma patients (**FIGURE** S6), with patients aged 70 or younger having a better survival than those patients older than 70 years (log-rank p-value = 0.008, median survival difference 6.9 months). There was no correlation between age and NHOCpeak (Spearman’s ρ = –9×10^-4^). Despite the prognostic value of both variables separately, the combination of both through a Cox regression model did not yield significant results (p-value = 0.088).

## DISCUSSION

The increasing availability of medical data is accelerating the development of personalised biomarkers (*24–26*). Nuclear and molecular imaging enable direct visualisation of cancer processes at the macroscopic level, making them powerful tools for assessing a patient’s disease state (*27,28*). At the same time, mathematical modelling is leading to an increasing number of identified biomarkers (*29,30*). The NHOC metric emerged from computational simulations and mathematical analysis indicating that metabolic activity shifts toward the tumour periphery as tumours progress. This led to the finding that NSCLC and breast cancers with more peripheral metabolic activity—higher NHOC—are associated with poorer survival outcomes (*13*).

Here, we demonstrate that this biomarker also holds prognostic value in a cohort of 61 patients with high-grade glioma imaged with ^18^F-FCHOL. In this case, NHOCpeak was used, defined as the relative distance between the SUVpeak and the tumour centroid, normalized by the mean spherical radius (*13,14*). A subset comprising one quarter of the patients with low NHOCpeak showed improved prognosis, with a 10.1-month median survival increase. It is striking that the same modeling framework succeeds across such different malignancies, despite their significantly different biological behaviours. This highlights the fundamental nature of the underlying mechanisms, namely the displacement of tumour proliferation and evolutionary dynamics into regions of available space.

Remarkably, it is the NHOCpeak that best selects the favourable cases in glioma, but not NHOCmax. Our analyses point to a higher spatial heterogeneity in the metabolic profiles for this group in which SUVpeak might be a better indicator of where the maximum activity is located. The absence of thoracic motion existing in previous cohorts, added to a smaller voxel size, might result in images with greater retained noise in glioma. Therefore, the process of filtering needed to compute the SUVpeak might actually help better identify the right location of the activity hotspot (*31–33*).

Another interesting difference of this high-grade glioma cohort is that values of NHOCmax and NHOCpeak are higher than most previously published groups (see **FIGURE S1**). The mean ± standard deviation of NHOCpeak here is 0.65 ± 0.25, while it was respectively 0.31 ± 0.2 and 0.37 ± 0.21 for the original NSCLC and breast cancer cohorts (*13*), and 0.54 ± 0.29 for an independently published NSCLC cohort (*15*). Therefore, the evidence here indicates that gliomas have their activity much less centred than NSCLC and breast cancers, which is consistent with their fast growth and the formation of an inner necrotic core.

Interestingly, the division between groups with different survival in our cohort happened at the middle between the two extreme cases (NHOCpeak = 0.495). A similar threshold for NHOCmax had been observed in the original breast cancer cohort (*13*)—threshold for NHOCmax = 0.499—and for the NSCLC group subject to immunotherapy in ref. (*15*)—threshold for NHOCpeak = 0.502. Deviations from this behaviour are the original NSCLC cohort—threshold for NHOCmax = 0.64—and the breast cancer cohort in ref. (*16*)—threshold for NHOCpeak = 0.27. Even though further research is still needed to understand the critical NHOC values across histologies, the critical value NHOC = 0.5 might get established as a general reference valid for many different cancers.

Beyond the use of NHOCpeak as a standalone biomarker, the combination with the SUVpeak revealed informative insights. Each patient was stratified into four subgroups according to their value of SUVpeak (low/high) and NHOCpeak (low/high). Patients with low SUVpeak and low NHOCpeak showed excellent survival, with a median survival increase of 16.4 months compared to the second-best group, and 21.9 months to the group with lower survival (high SUVpeak/high NHOCpeak). Therefore, within the patients with low SUVpeak, the value of NHOCpeak significantly stratified survival outcomes, while patients with both low NHOCpeak and low SUVpeak had markedly increased survival.

The distribution of points in **FIGURE 4**(A) and their associated survival outcomes (**FIGURE 4**(B)) suggest a potenatial disease evolutionary trajectory: from low SUVpeak and low NHOCpeak (Group 1, excellent survival) to high NHOCpeak and low SUVpeak (Group 2, intermediate survival), followed by an increase in metabolic activity (Group 3). In agreement with this trajectory, few patients exhibit high SUVpeak with low NHOCpeak (Group 4). Following this hypothesis, NHOCpeak would distinguish markedly different stages of the disease and SUVpeak would provide additional prognostic resolution in already advanced cases.

Despite its findings, this study has limitations. The most important one is that the cohort studied here includes high-grade gliomas from different histologies, including both IDH-WT, classical glioblastomas, high-grade anaplastic astrocytomas, and one case of oligodendroglioma. This is a consequence of the retrospective nature of the study, which collected patients along the years and whose beginning dates to older glioma classification (*34*). Future prospective studies should consider the modern classification and consider the relationships with molecular biomarkers (*35*). We examined whether the values of the NHOCpeak might be related to different IDH status but obtained inconclusive results. On the one hand, the relationship between NHOCpeak groups did not significantly associate with the IDH status, but on the other hand the biomarker was not successful when IDH-mut cases were removed from the cohort. Larger cohorts will be needed to clarify these relationships. Moreover, the use of other PET radiotracers such as ^18^F-FDOPA will enable the evaluation of NHOC basis and performance under different metabolic conditions (*36,37*).

## CONCLUSION

The obtained biomarker NHOCpeak, which defines the location of the voxel of SUVpeak in the tumour centroid-edge axis, contains prognosis information about survival in high-grade gliomas. In particular, for patients with low SUVpeak on ^18^F-FCHOL PET, the NHOCpeak value distinguished clear differences in survival.

## Data Availability

All data produced in the present study are available upon reasonable request to the authors

## Supplementary Information

**FIGURE S1.**
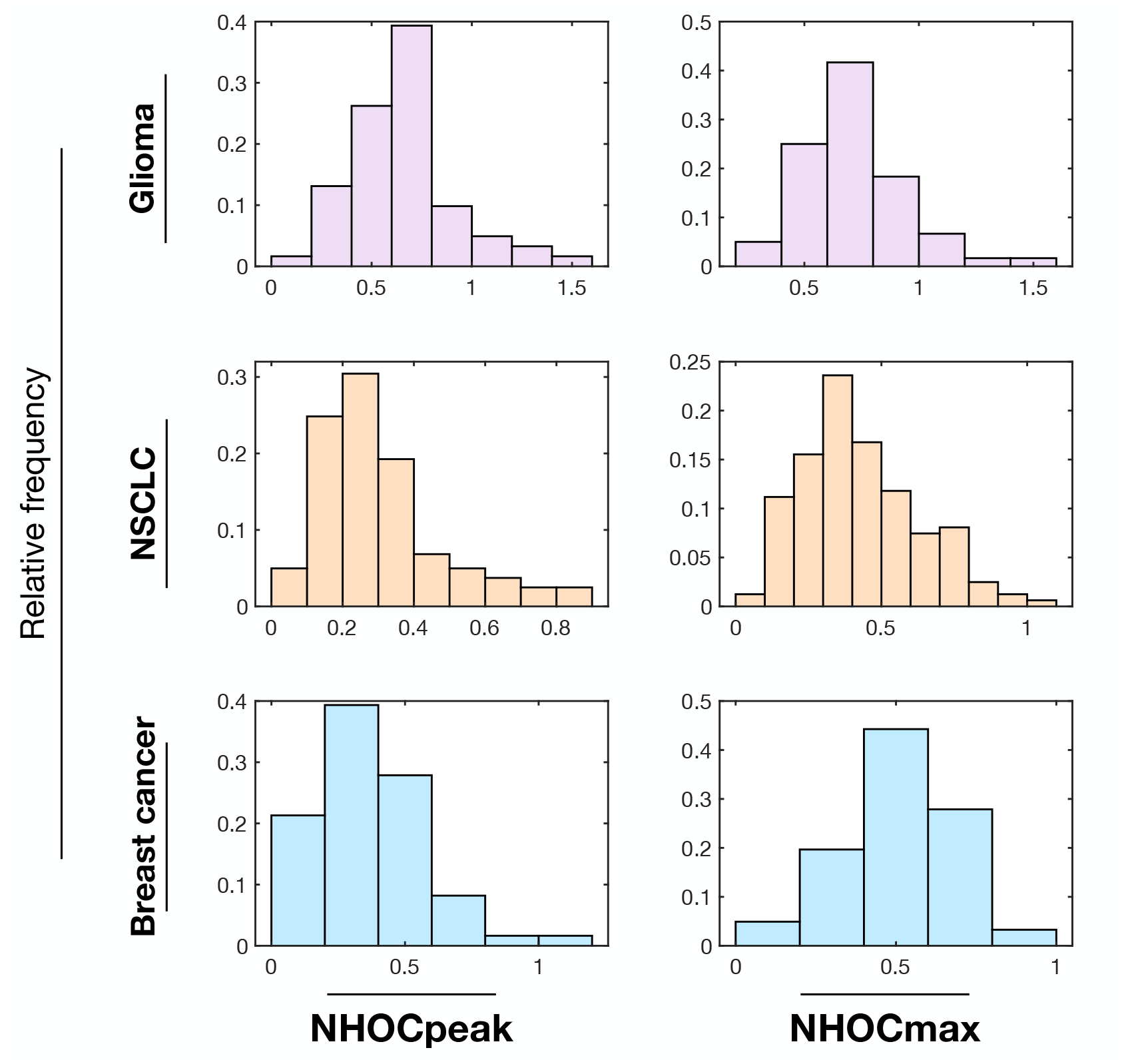
Histograms for the metrics NHOCpeak and NHOCmax in patients with high-grade glioma (first row) and patients with NSCLC and breast cancer (see Materials and Methods).

**TABLE S1.** Contingency table for patients with known IDH status.

|  | IDH-WT | IDH-mut | Row total |
| --- | --- | --- | --- |
| **NHOCpeak > 0.495** | 36 (62 %) | 7 (12 %) | 43 (74 %) |
| **NHOCpeak ≤ 0.495** | 10 (17 %) | 5 (9 %) | 15 (26 %) |
| Column total | 46 (79 %) | 12 (21 %) | 58 (100 %) |

**FIGURE S2.**
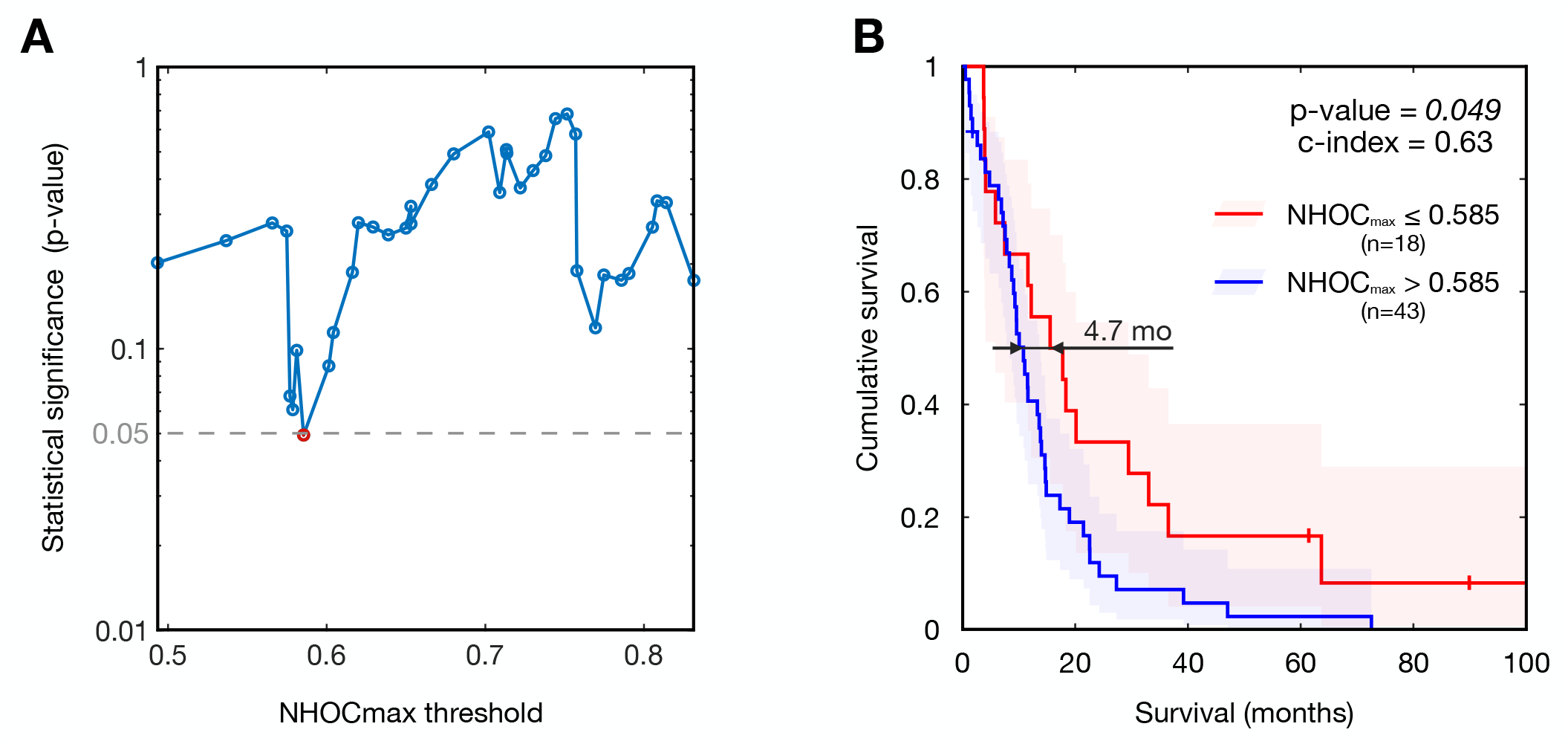
NHOCpeak survival analysis. (A) Log rank p-values obtained from the survival analysis of the two group separations by variable thresholds (x axis). (B) Kaplan–Meier curve corresponding to the NHOCpeak that separates the patient’s groups with the best equilibrium between significance level and equilibrium between patients in each group.

**FIGURE S3.**
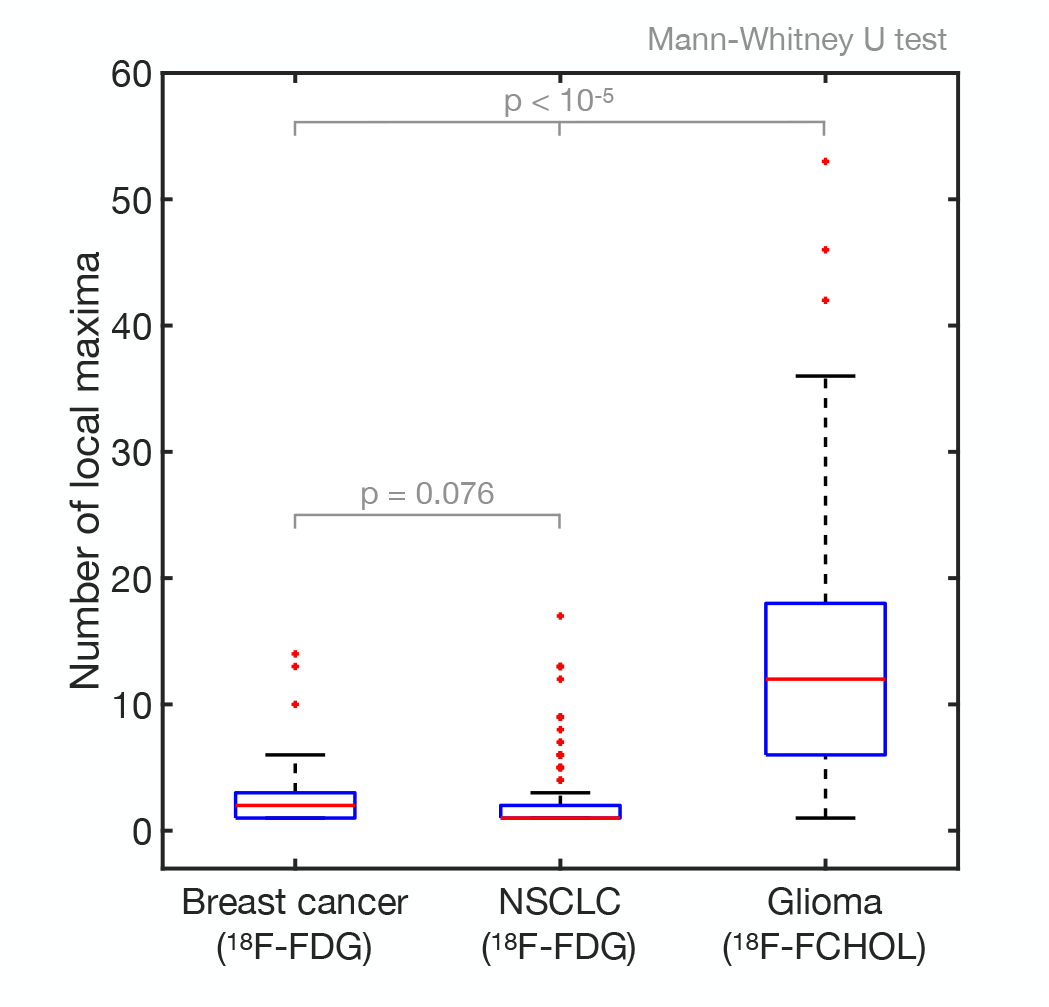
Box plots for number of local maxima (voxels with a higher activity than their surroundings) existing in the delineated tumours.

**FIGURE S4.**
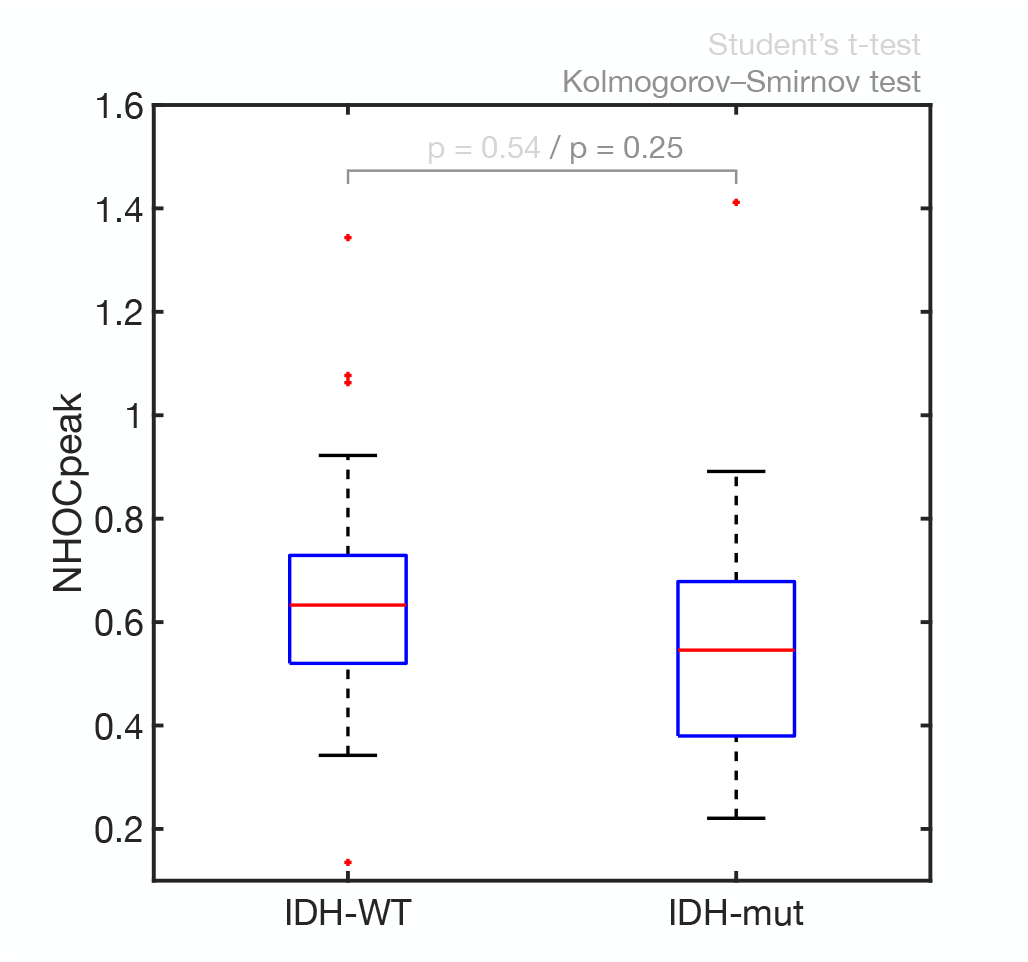
NHOCpeak box plots for patients with IDH-WT and patients with IDH-mut. Statistical tests do not reflect differences between groups.

**FIGURE S5.**
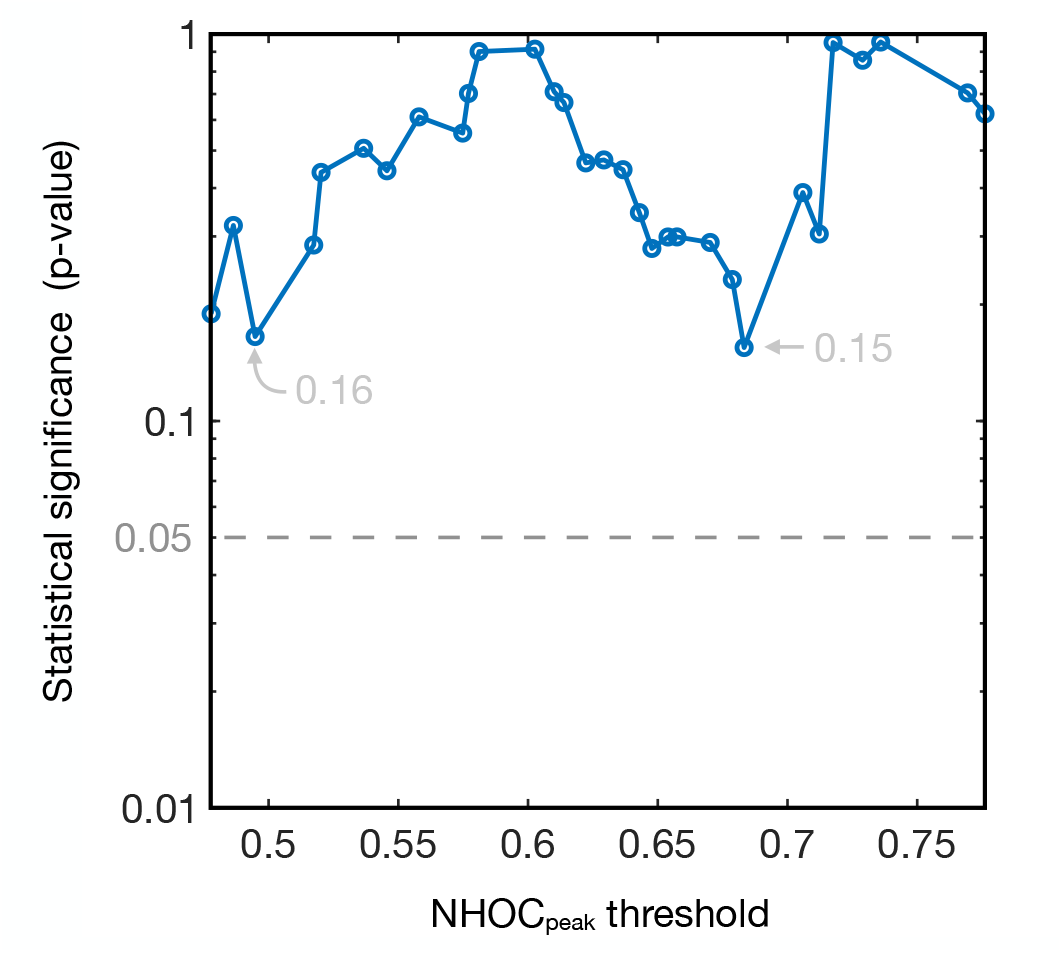
NHOCpeak survival analysis for patients with IDH-WT status. The figure shows the log rank p-values obtained from the survival analysis of the two group separations by variable thresholds (x axis).

**TABLE S2.** Spearman’s correlation coefficients between PET biomarkers and MRI biomarkers.

|  | **Total volume** | **Necrotic volume** | **Contrast enhancement**<br>**rim width** | **Tumour**<br>**surface** | **Surface**<br>**regularity** |
| --- | --- | --- | --- | --- | --- |
| **NHOCpeak** | 0.055 | 0.185 | -0.047 | 0.087 | 0.077 |
| **SUVpeak** | 0.086 | -0.019 | 0.175 | 0.099 | 0.058 |

**FIGURE S6.**
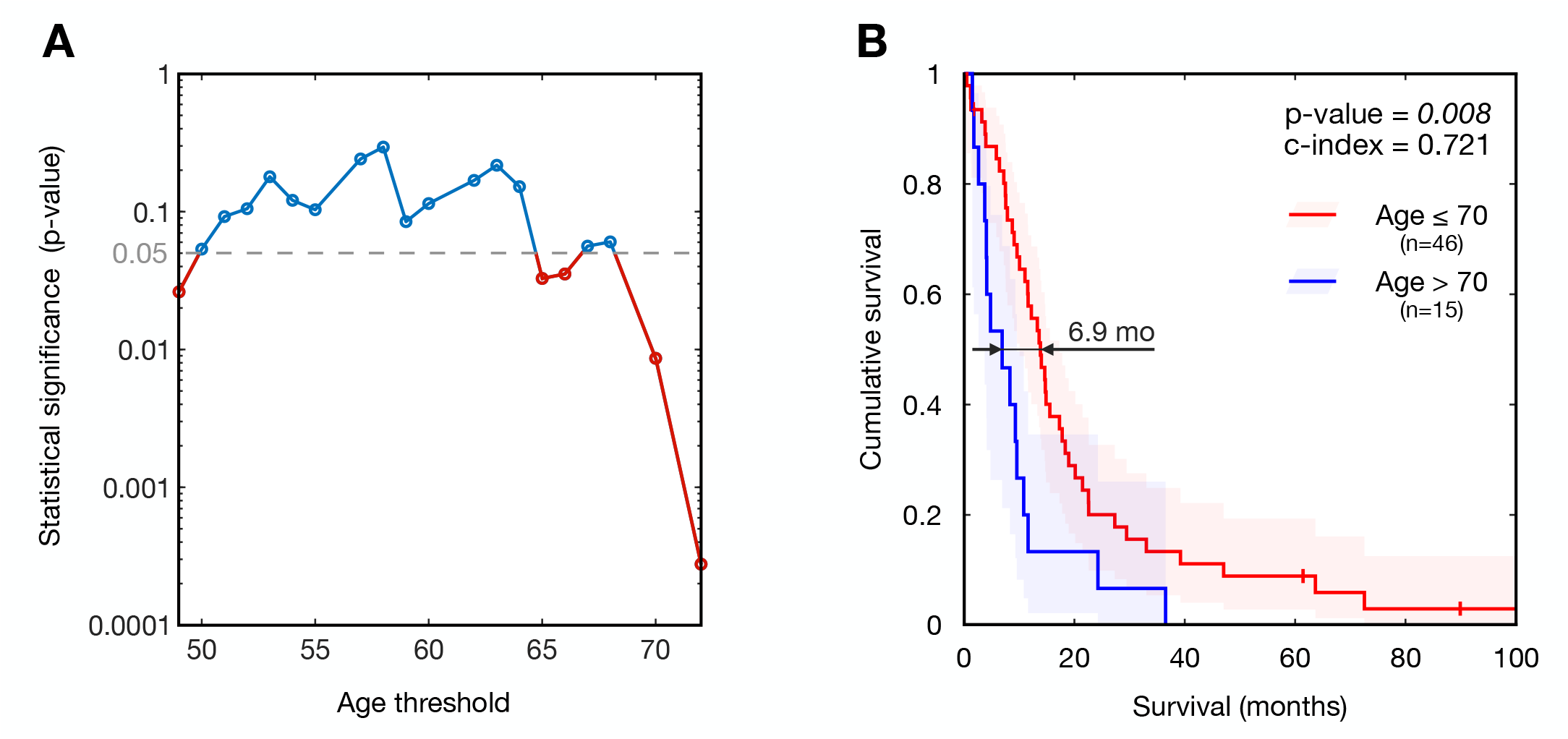
Age survival analysis. (A) Log rank p-values obtained from the survival analysis of the two group separations by variable thresholds (x axis). (B) Kaplan–Meier curve corresponding to the 70 years-old.

